# Insights from a Pan India Sero-Epidemiological survey (Phenome-India Cohort) for SARS-CoV-2

**DOI:** 10.1101/2021.01.12.21249713

**Authors:** Salwa Naushin, Viren Sardana, Rajat Ujjainiya, Nitin Bhatheja, Rintu Kutum, Akash Kumar Bhaskar, Shalini Pradhan, Satyartha Prakash, Raju Khan, Birendra Singh Rawat, Karthik Bharadwaj Tallapaka, Mahesh Anumalla, Giriraj Ratan Chandak, Amit Lahiri, Susanta Kar, Shrikant Ramesh Mulay, Madhav Nilakanth Mugale, Mrigank Srivastava, Shaziya Khan, Anjali Srivastava, Bhawna Tomar, Murugan Veerapandian, Ganesh Venkatachalam, Selvamani Raja Vijayakumar, Ajay Agarwal, Dinesh Gupta, Prakash M Halami, Muthukumar Serva Peddha, Gopinath M Sundaram, Ravindra P Veeranna, Anirban Pal, Vinay Kumar Agarwal, Anil Ku Maurya, Ran Vijay Kumar Singh, Ashok Kumar Raman, Suresh Kumar Anandasadagopan, Parimala Karuppanan, Subramanian Venkatesan, Harish Kumar Sardana, Anamika Kothari, Rishabh Jain, Anupma Thakur, Devendra Singh Parihar, Anas Saifi, Jasleen Kaur, Virendra Kumar, Avinash Mishra, Iranna Goger, Geethavani Rayasam, Praveen Singh, Rahul Chakraborty, Gaura Chaturvedi, Pinreddy Karunakar, Rohit Yadav, Sunanda Singhmar, Dayanidhi Singh, Sharmistha Sarkar, Purbasha Bhattacharya, Sundaram Acharya, Vandana Singh, Shweta Verma, Drishti Soni, Surabhi Seth, Shakshi Vashisht, Sarita Thakran, Firdaus Fatima, Akash Pratap Singh, Akanksha Sharma, Babita Sharma, Manikandan Subramanian, Yogendra Padwad, Vipin Hallan, Vikram Patial, Damanpreet Singh, Narendra Vijay Tirpude, Partha Chakrabarti, Sujay Krishna Maity, Dipyaman Ganguly, Jit Sarkar, Sistla Ramakrishna, Balthu Narender Kumar, A Kiran Kumar, Sumit G. Gandhi, Piyush Singh Jamwal, Rekha Chouhan, Vijay Lakshmi Jamwal, Nitika Kapoor, Debashish Ghosh, Ghanshyam Thakkar, Umakanta Subudhi, Pradip Sen, Saumya Ray Chaudhury, Rashmi Kumar, Pawan Gupta, Amit Tuli, Deepak Sharma, Rajesh P. Ringe, Amarnarayan D, Mahesh Kulkarni, Dhanasekaran Shanmugam, Mahesh S Dharne, Syed G. Dastager, Rakesh Joshi, Amita P. Patil, Sachin N. Mahajan, Abu Junaid Khan, Vasudev Wagh, Rakeshkumar Yadav, Ajinkya Khilari, Mayuri Bhadange, Arvindkumar H. Chaurasiya, Shabda E Kulsange, Krishna Khairnar, Shilpa Paranjape, Jatin Kalita, G. Narahari Sastry, Tridip Phukan, Prasenjit Manna, Wahengbam Romi, Pankaj Bharali, Dibyajyoti Ozah, Ravi Kumar Sahu, Elapavalooru V.S.S.K. Babu, Rajeev Sukumaran, Aiswarya R Nair, Prajeesh Kooloth-Valappil, Anoop Puthiyamadam, Adarsh Velayudhanpillai, Kalpana Chodankar, Samir Damare, Yennapu Madhavi, Ved Varun Aggarwal, Sumit Dahiya, Anurag Agrawal, Debasis Dash, Shantanu Sengupta

## Abstract

To understand the spread of SARS-CoV2, in August and September 2020, the Council of Scientific and Industrial Research (India), conducted a sero-survey across its constituent laboratories and centers across India. Of 10,427 volunteers, 1058 (10.14%) tested positive for SARS CoV2 anti-nucleocapsid (anti-NC) antibodies; 95% with surrogate neutralization activity. Three-fourth recalled no symptoms. Repeat serology tests at 3 (n=346) and 6 (n=35) months confirmed stability of antibody response and neutralization potential. Local sero-positivity was higher in densely populated cities and was inversely correlated with a 30 day change in regional test positivity rates (TPR). Regional seropositivity above 10% was associated with declining TPR. Personal factors associated with higher odds of sero-positivity were high-exposure work (Odds Ratio, 95% CI, p value; 2·23, 1·92–2·59, 6·5E-26), use of public transport (1·79, 1·43–2·24, 2·8E-06), not smoking (1·52, 1·16–1·99, 0·02), non-vegetarian diet (1·67, 1·41–1·99, 3·0E-08), and B blood group (1·36,1·15-1·61, 0·001).

**Impact Statement:** Widespread asymptomatic and undetected SARS-CoV2 infection affected more than a 100 million Indians by September 2020. Declining new cases thereafter may be due to persisting humoral immunity amongst sub-communities with high exposure.

**Funding:** Council of Scientific and Industrial Research, India (CSIR)

## Introduction

The World Health Organization declared SARS-CoV-2 infection as a pandemic on March 11 2020.^1^ Within two weeks, India announced a lockdown strategy that severely influenced the growth of the pandemic which was initially very focal in the large cities, gathering pace and spreading to smaller cities and towns as the nation unlocked for societal and economic considerations.

Early literature pointed towards asymptomatic transmission of SARS-CoV-2 and raised the need for extended testing.^2,3^ While, RT-PCR was an undisputed choice for establishing a positive infection, sero-surveillance revealed that many more were probably getting infected without manifesting symptoms.^4,5^ Initial estimates of asymptomatic infection rate from the West were around 40-45%.^5^

In India, the first case of Covid was reported on January 30, 2020.^6^ Serological surveys have confirmed that spread beyond the Indian megacities was minimal in early May-June, with less than 1% sero-positivity outside the designated containment zones, suggesting that the lockdown had been effective in limiting the spread.^7^ This was not without human and economic cost. By the end of June, migrant workforces caught in the cities during the lockdown were sent to their rural homes, which may have led to subsequent rapid, multi-focal rise in cases in July 2020. By October 2020, total new cases began to decline with further outbreaks limited to a few geographies. Thus the period of August to September 2020 is an important transition point. Existing studies from India, have been limited to specific geographies or localities.^8-10^ The present study is one of two studies at a national scale that was designed to assess spread of infection. A national sero-survey in 70 districts of India conducted by ICMR, is yet to be published, but, had a reported crude positivity rate of about 10%.^11^ The current study was launched by the Council of Scientific and Industrial Research (CSIR) in its more than forty constituent laboratories and centers spread all over the country, representing a wide range of ethnicities, geo-social habitats and occupational exposures, in the form of a longitudinal cohort (Phenome-India Cohort). While limited by not being a population denominated study, the cohort includes permanent staff, families of staff members, students, and temporary employees proving support services such as security, sanitation, housekeeping etc. This is a diverse microcosm of India encompassing multiple socio-economic groups and has the advantage of permitting deeper assessments such as questionnaire surveys and periodic reassessment of humoral antibody response in those found to be sero-positive. Here, we report results from phase 1 of this study covering the critical period of August to September 2020.

## Methods

### Study Design, Sampling and Data Collection

The longitudinal cohort study was approved by Institutional ethics committee of CSIR-IGIB. The minimal sample size for estimating seropositivity of about 5% with 10 percent precision (0.005) with 95% confidence was 7300.^12^ In the current study, we enrolled greater than 10000 subjects. 10427 adult subjects working in CSIR laboratories and their family members enrolled for the study based on voluntary participation. Informed consent was obtained from all the participants and the samples were collected maintaining all recommended precautions. Blood samples (6 ml) were collected in EDTA vials from each participant and antibodies to SARS-CoV-m 2 nucleocapsid antigen were measured using an Electro-chemiluminescence Immunoassay (ECLIA)-Elecsys Anti-SARS-CoV-2 kit (Roche Diagnostics) as per the manufacturer’s protocol. This approved assay s considered a method of choice when a single test is to be deployed.^13^ A COI ≥ 1 was considered sero-positive. Positive samples were further tested for neutralizing antibody (NAB) response directed against the spike protein using GENScript cPass™ SARS-CoV-2 Neutralization Antibody Detection Kit (Genscript, USA), according to the manufacturer’s protocol. This is a blocking ELISA used for qualitative detection of total neutralizing antibodies against SARS-CoV-2 virus in plasma. A value of 20% or above was considered to have neutralizing ability.

All the participants were requested to fill an online questionnaire, which included information on date of birth, gender, blood group, type of occupation, history of Diabetes, Hypertension, Cardiovascular Disease, Liver and Kidney Disease, diet preferences, mode of travel, contact history, and hospital visits. These forms were then downloaded in MS-Excel data format and merged with registration forms filled at the time of sample collection based on unique ID’s.

### Data and Statistical Analysis

Region wise and total sero-positivity was calculated from the fraction of samples positive for antibodies to SARS-CoV-2 nucleocapsid antigen. Data regarding rtPCR/rapid antigen testing and positive cases was gathered from the website “covid19india.org”. Change in Test Positivity Rate (TPR); a robust parameter for estimating the level of infection transmission when level of testing is variable, was calculated as per following equation:

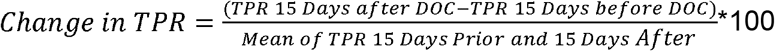

#### *DOC-Date of Collection

IGIB, New Delhi and NAL, Bengaluru were removed from this Change in TPR analysis, for, the samples collection was spread over 2-3 weeks in these labs. Questionnaire based variables were assessed for responses type and blank fields i.e. responses which were not provided by the participants of the survey. Based on multiple response types for each variable, categories were made to assign the response to either of the categories. For visualization; ggpubr (v0.4.0), ggrepel (v0.8.2), ggplot2 (v3.3.2) packages were used in R. No data imputation was carried out. Chi-square test was performed to evaluate variables which had significant association with outcome of being tested positive (p<0·05) along with Odds Ratio (OR) with 95% Confidence Interval (CI). An adjusted p value was obtained through Bonferroni Correction method for multiple comparison testing. Following the chi-square test an iterative logistic regression was carried out on balanced dataset. Variance Inflation Factor (VIF) was separately evaluated to assess multi-collinearity. Statistical analysis and model development was carried out with visualization in R programing environment version 3.6.1, MS-Excel 2016 and OriginPro V2021; faraway (v1.0.7) package was utilized for estimation of VIF.

## Results

### Seropositivity varied widely across India

In 10427 subjects from over 17 States and two Union Territories, the average sero-positivity was 10·14% (95% CI 9.6-10.7), but varied widely across locations (Figure 1). We found that 95% of the sero-positive subjects also had significant neutralizing activity, suggesting at least partial immunity (Figure 1 Source Data).

**Figure 1:**
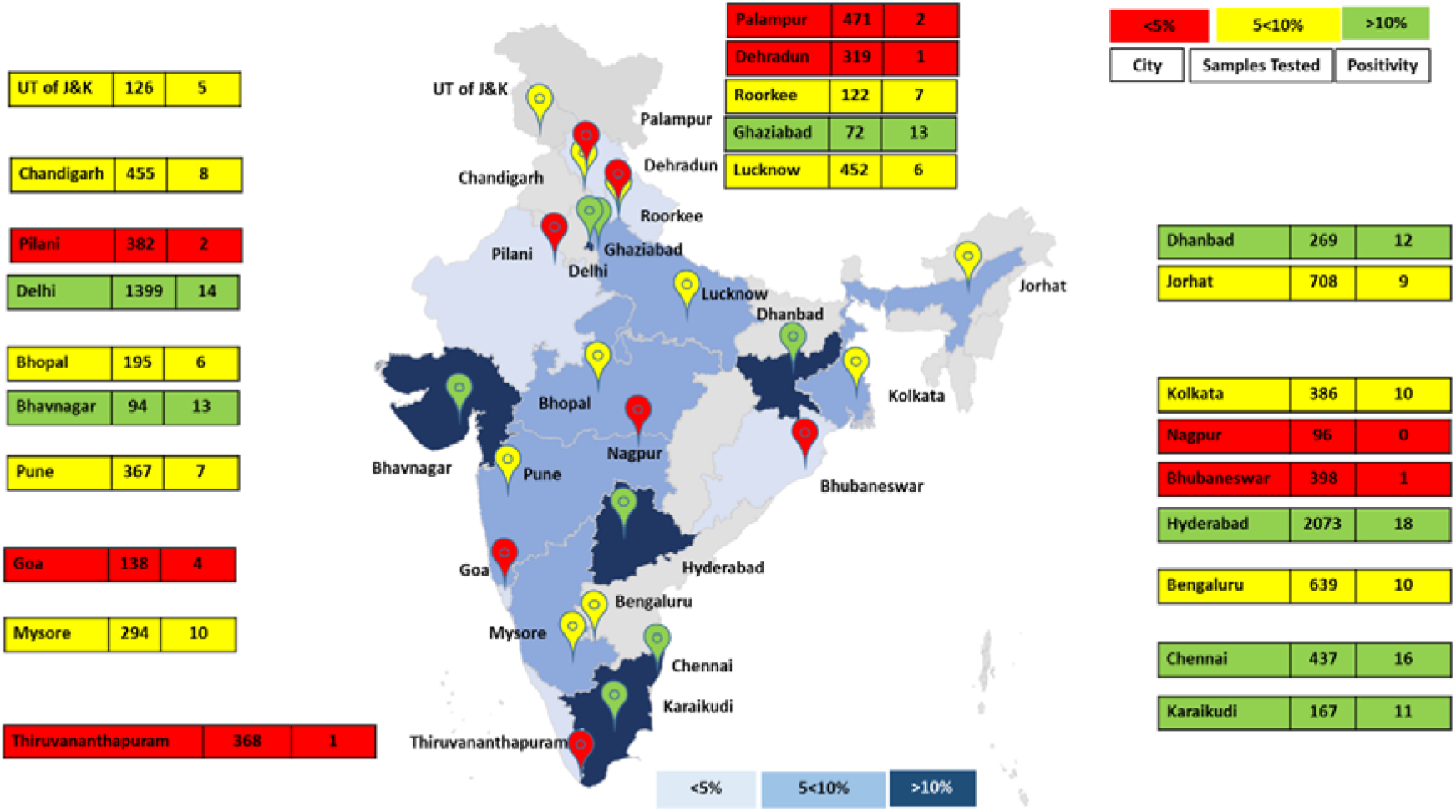
City wise total samples collected and sero-positivity (India map may not be to scale and is for representation purposes only and Sero-Positivity is rounded off).

### Sero-positivity, population density, and trajectory of new infections

As expected from the known outward spread of infection from large Indian cities, seropositivity was greater in regions with higher population density (Figure 2A). Lab-wise seropositivity was correlated with the regional change in Test Positivity rate (TPR). Changes in TPR are a relatively robust marker of the local level of transmission and are preferred when absolute number of tests or test rates are variable, as was the case here. By this measure, regional transmission of SARS-CoV2 was inversely correlated to local seropositivity. Seropositivity of 10% or more was associated with reductions in TPR which may mean declining transmission (Figure 2B and Figure 2B Source Data).

**Figure 2:**
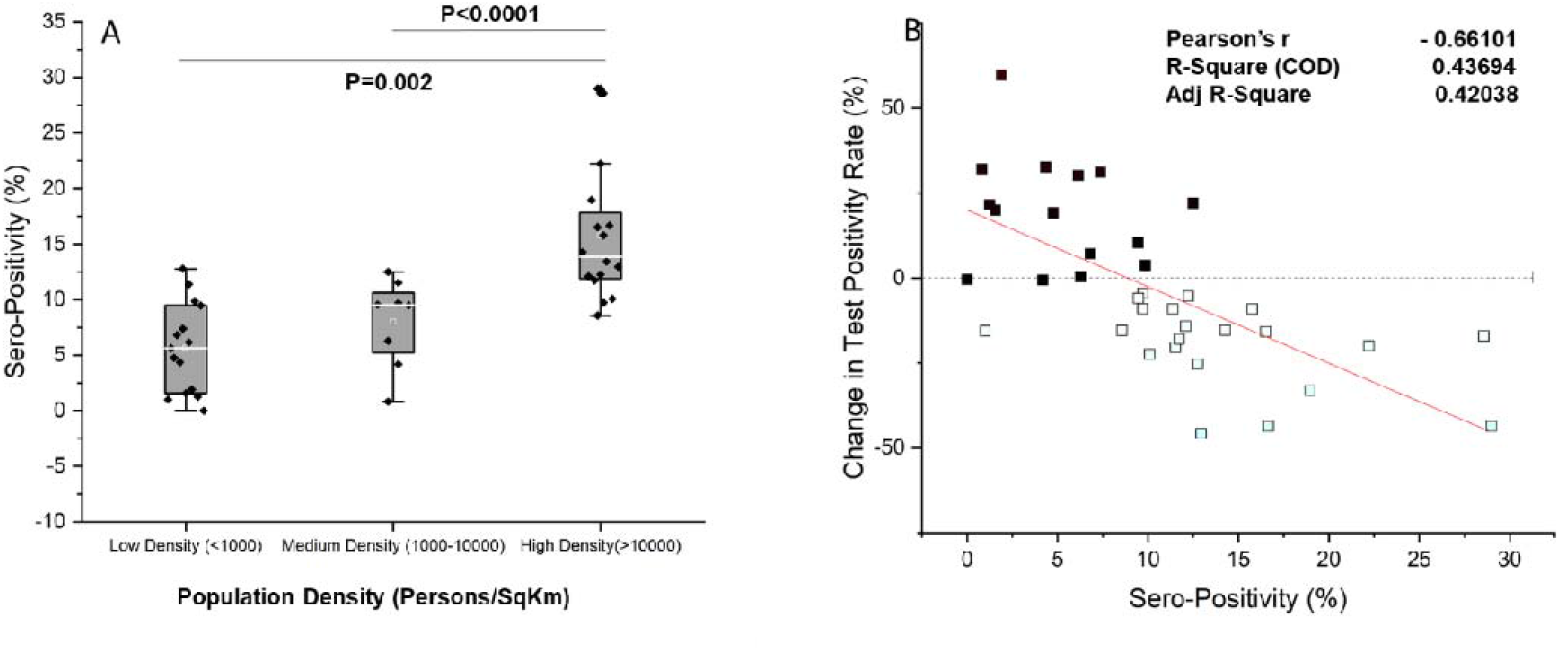
A) Population density based sero-positivity (y-axis) for labs/centers samples (x-axis), an overall p value of <0.0001 was obtained on one way ANOVA. B) Change in Test Positivity Rate (%) for stae COVID 19 infections (y-axis) against observed sero-positivity of labs/centers (x-axis). A negative slope reflects declining TPR with increase in sero-positivity.

*Figure 1 and Figure 2 Source Data: Data for all labs/centers utilized for Figure 2A and Figure 2B. Lab, District and Date of Collection (DOC) [Column A-C] Total Samples Collected, Number of sero-positive samples, sero-positivity in percentage (rounded Off), Number of samples tested and found positive for NAb. Columns [E-I] Number of confirmed cases and tests done for respective states 15 days and prior and after the DOC. Columns [L-N] Data obtained from *www.covid19India.org*) (State Data has been utilized as a surrogate to City/ District Data for City/District data was not available for number of cases/tests done for many)*.

### Survey based correlates of seropositivity

Using seropositivity as a surrogate of prior infection, the survey data enabled exploration of the typical clinical course as well as risk-factors for infection.

Out of 861 sero-positive subjects who also provided data on symptomatology, 647 subjects (75·3%) did not recall any of the nine symptoms asked for. Amongst the minority of subjects with symptoms, the most commonly reported symptom constellation was those of a mild flu-like disease with fever (∼50 %) as the most frequent symptom. Loss of smell or taste was uncommonly reported (∼25% of symptomatic subjects). An observation was also made; 19 percent amongst asymptomatic sero-positives were females, it nearly doubled to 36 percent amongst symptomatic sero-positives.

We further examined associations of other available variables with sero-positivity to explore potential factors that modulate risk of infection in India. Apart from gender and age, distribution of the other variables recorded in CSIR-cohort (prevalence of smoking, diet, physiological parameters like ABO blood group type) were similar to the national averages and the sample can be considered representative.^14,15^ The associations are shown in Table 1, separately for each gender as well as combined.

**Table 1.**
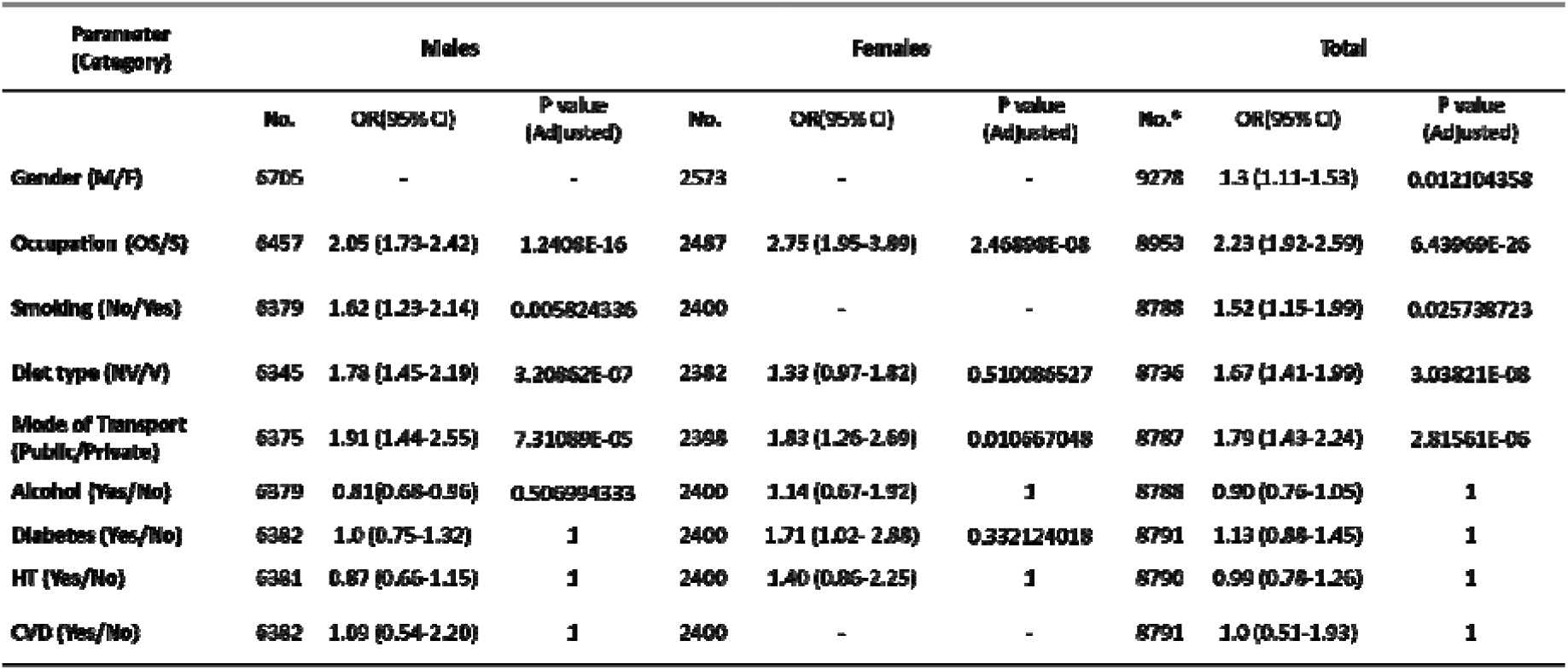
Demographics of data available for different variables (M- Males, F- Females, OS-Outsourced Staff, S-Staff, NV-Non-Vegetarian, V-Vegetarian, HT-Hypertension, CVD-Cardiovascular Disease, OR-Odds Ratio, CI-Confidence interval. *Male and female numbers not adding to total as gender field was not available or filled by these participants)

Gender distribution in our dataset was highly unequal (72% males and 28% females). While we observed differences in sero-positivity by gender, 10·43% among males, vs 8·20% among females (OR 1·30 95% CI 1·11 -1·53, p=0·012), this association was not significant when tested in an iteration model with balanced dataset.

Blood Group type was reported for 7496 subjects. Blood Group (BG) distribution of subjects in our study was similar to national reference based on a recent systematic review.^14^ Sero-positivity was highest for blood group type AB (10·19%) followed by B (9·94%), O (7·09%) and A (6·97%). Blood O may be associated with a lower sero-positivity rate, with an OR of 0·76 (95 % CI 0·64 -0·91, p=0·018) vs Non O blood group types, while B appeared to be high-risk with an OR of 1·36 (95 % CI 1·15 -1·61, p=0·001). Blood group A had an OR of 0.78, the association was not found to be significant (p=0.10), while a similar observation was made with blood group AB (p=0.35), it had an OR of 1.27. Rh factor was not found to have significant association with sero-positivity (p=0·35).

Occupational risks and lifestyle were important determinants of risk as shown in Table 1. Some of these were along expected lines of higher exposure such as high-contact occupations of outsourced staff or the use of public transport. Interestingly, personal habits such as smoking or vegetarian diet were observed to be associated with lower sero-positivity. Non-smokers in the cohort recorded a significantly higher sero-positivity of 10·11%, as opposed to smokers (6·88%). Non-vegetarians had a sero-prevalence of 11%, while sero-positivity among vegetarians was 6·86%. The results for odds ratio and iteratively run regression model for entire dataset and separately for male and female gender are shown in Figure 3. Presence or absence of Diabetes, Hypertension and Cardiovascular Disease were not found to be significantly associated with sero-outcomes (p>0·05, Table 1).

**Figure 3.**
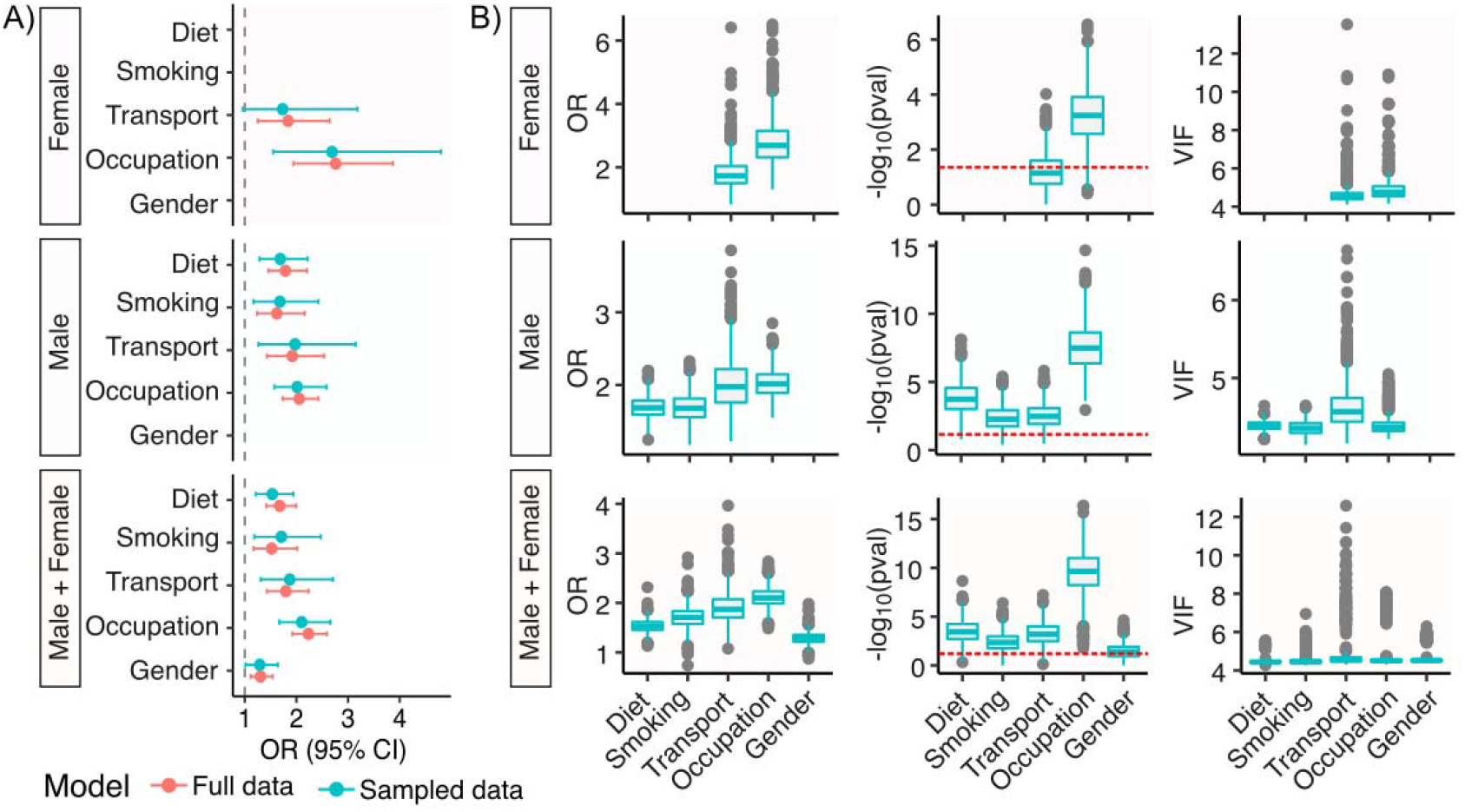
A) Odds Ratio of full dataset and sampled data set obtained from model. B) Odds, p value and VIF for sampled dataset with iterations on regression model. (For Diet: Non-Vegetarian against Vegetarian, For Smoking: Non-Smoking against Smoking, For Transportation: Public against Private, For Occupation: Outsourced Staff against Staff, For Gender: Male against Female). 1300, 1100 and 1000 iterations were run for Female, Male +Female and Males respectively.

### Stability of humoral response to SARS-CoV-2

Of 346 subjects whose samples were collected again at three months, anti-nucleocapsid antibody levels were similar or higher for most, with only five (1·4%) becoming sero-negative (Figure 4A). In contrast, neutralizing activity was lost (below 20%) in 11 subjects (Figure 4B). However, in 35 subjects who could be further re-tested at six months; the anti-nucleocapsid antibody levels declined while neutralizing antibody levels were mostly unaltered when compared to the 3 months level (Figure 4C and D).

**Figure 4:**
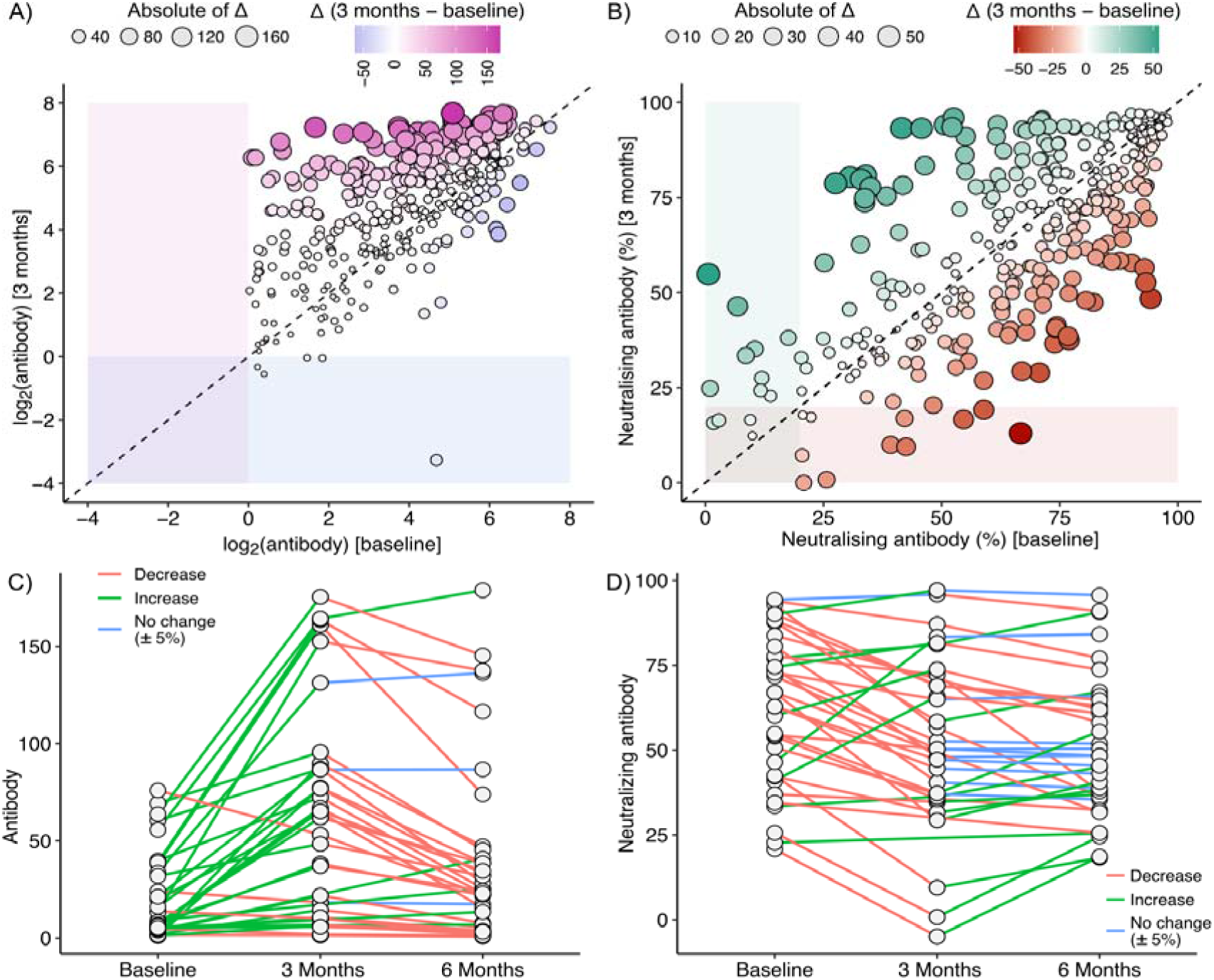
Antibody levels (A) and Neutralizing Antibody percentage (B) level at baseline (x-axis) and after 3 months (y-axis). Antibody levels (C) and Neutralizing Antibody (D) percentage level (y-axis) at baseline, 3 months and after 6 months (x-axis) for 35 subjects depicting the trend.

## Discussion

The present study, which recruited subjects from 24 cities in India, provides an important and timely snapshot of the spread of SARS CoV2 pandemic across India shortly before the peak of new cases. It confirms that by September 2020, a large pool of recovered Indians with at least partial immunity existed. Between our study and unpublished numbers from a national sero-survey at the same time, more than a hundred million Indians were likely to belong to this category. The subsequent nationwide decline of new cases starting in October 2020 can be well understood through these observations. There is some evidence of declining transmission in high seropositivity regions within September, based on falling test positivity rates, but due to changes in tests and expansion of testing, this can only be indirectly inferred. As shown in our study, the fraction of such recovered subjects with resistance to reinfection was more than double amongst people performing high-contact jobs and using public transport. Thus, it is not surprising that in combination with a strong emphasis on masking and distancing, new cases started declining soon after this sero-survey. As of January 2021, despite the onset of winter and new year festivities, India has not seen major outbreaks. It is important to note that the crude sero-positivity rate reported by us needs adjustments for demographics, fraction of infected subjects who may not develop antibodies, test characteristics, amongst others, to be a true population seropositivity rate. Here, we intentionally avoid such adjustments since it provides a false sense of precision with too many unknowns and un-measurables. We focus on the more meaningful variation of the crude rate, its clinical correlates, and public health implications. We are confident from the data that a very large pool of recovered and immune subjects existed by September 2020, as stated, and expect that downward adjustments for national demographics will be counterbalanced by upward adjustments for almost 20% of rtPCR proven asymptomatic infections who develop transient antibody responses.^16^

Apart from the sero-positivity rate, our data also reveals important associations between demographic, physiological, lifestyle-related and occupational attributes with susceptibility to infection. The workforce in our cohort comprised of adult population and no major difference was observed in sero-positivity amongst different age groups or those with co-morbidities. Males were found to be more susceptible, in agreement with other published reports^17^. However, there were fewer females in our study and many of the occupational responsibilities with higher chances of exposure, like that of security personnel, were skewed towards males. On iteratively ran regression models we did not find gender to be a predictor for sero-positivity.

ABO blood group type has been shown to be associated with SARS-CoV-2 infection, but the results are variable in different studies. Most studies found that O group is associated with lower risk of infection or severity and Blood Group A was reported to be high risk in some studies^18-21^. In a meta-analysis authored by Golinelli et al^22^, they observed positive association with A blood group, while blood group O was to be associated with lesser positivity using a random effects model. Another study from India observed blood group O to be associated with less mortality while blood group B with higher mortality when they analyzed the national data available^23^. A complex molecular interaction is said to play a significant role, and the molecular pathways for the same need to be elucidated for the effects observed specially with blood group O, which was also confirmed by our study.

In regard to diet preferences, while an association was observed overall and in males, it needs to be corroborated with further elaborative studies. It has been proposed that a fiber-rich diet may play an important role in COVID-19 through anti-inflammatory properties by modification of gut microbiota.^24^ Vegetarian diet is known to have high fiber content and possibly may effect through microbial alteration, but an implied effect on immune-biology and lung biology is not yet elucidated.^24,25^ A recent review highlighted the role of trace elements, nutraceuticals and probiotics in COVID-19.^26^ These, through their immune-modulatory property exert an anti-viral effect. However, these observations should not advocate the usage or restriction of any diet type.

Our finding that smokers are less likely to be sero-positive is the first report from general population and part of growing evidence that despite COVID-19 being a respiratory disease, smoking may be associated with lower sero-positivity, though this association has not proven to be causal. Two studies from France and similar reports from Italy, New York and China reported lower infection rate among smokers.^27-31^ A Bayesian meta-analysis based review in early phase of the pandemic also reported reduced infection in current smokers.^32^ While, it was said the ACE 2 expression is higher and favorable to virus entry in smokers, increased mucous production through goblet cells may be acting as a first line of defense.^33,34^ Effect of increased nicotine receptor expression was also questioned.^33,35^ Hence, there is a need for focused mechanistic studies to understand the effect of smoking and nicotine on SARS-CoV-2 infection. Smoking is known to be severely detrimental to health and associated with multiple diseases and this observation should not be taken to be an endorsement, especially given that the association is not proved to be causal.

Serial follow up of antibody response provides important insights and there are limited studies published as of date in regard to it.^36-38^, specially from India. The persistence of neutralizing antibodies in our studies suggests that there may be immunity lasting for at least six months.

## Conclusion

The aggregate sero-positivity of 10·14% in our multi-centric study with diverse participants, with stable neutralizing activity that lasts for at least 6 months, suggests India had a large pool of recovered immune subjects by September 2020, especially amongst its high contact workers and people using public transport.

## Data Availability

Deidentified categorized data of the participants can be provided on request basis after approval of ethics committee of the institute/organisation to which the requesting person belongs, stating the potential need and usage and signing of an agreement with the corresponding authors on behalf of CSIR IGIB. The request could be made to the corresponding authors after 90 days of publication.

## Author Contribution

SSG, DD and AA conceptualized the idea and coordinated the research activity. SN, SSG, RajatU, AKB and SPradhan did all the sample analysis, storage and neutralization antibody experiments. VS, SSG, DD, NB, SN, RK, SPrakash, and RajatU contributed in Data Acquisition, Data Pre-Processing, Data Analysis. VS, RK, DD, SSG, and NB did statistical analysis and model development. RK, VS, SSG, DD and VKumar contributed in visualization and dashboard for analysis. RKhan, BSR, TKB, MA, GRC, AL, SKar, SRM, MNM, MS, SK, AS, BT, MV, GV, SRV, AjayA, DGupta, PMH, SPM, GM, PVR, APal, VKAgarwal, AKMaurya, RVKS, AKRaman, SRA, PK, SV, HKS, AKothari, AThakur, DSParihar, ASaifi, RJain, JKaur, AMishra, AmarD, IG, ASingh, PSingh, RC, GC, PKK, RYadav, DSingh, SS, SSarkar, PB, SA, VSingh, SVerma, DSoni, SSeth, SVashisht, ST, FF, APS, ASharma, BS, YP, VHallan, VPatial, DamanS, NVT, PC, SKM, DGanguly, JSarkar, SR, BNKumar, AKKumar, SGG, PSJ, RChouhan, VLJ, NK, DG, GT, US, PS, SRC, RKumar, PG, AT, DSharma, RPRinge, MK, DS, MDharne, SGD, RJoshi, APP, SNM, AJK, VW, RY, AK, MB, AHC, SEK, KK, SP, JK, GNS, TP, PM, WR, PBharali, DO, RKSahu, EB, RKS, AN, PKV, AP, AVP, KC, SD, YM, VVA, SDahiya, GR contributed in lab/ centre co-ordination, enrolment of volunteers, sample collection and logistics. VS, SSG, DD and AA wrote the first draft. SSG, VS, DD, DGanguly, GRC, Manikandan and AA critically revised the manuscript. All the authors agreed and contributed to the final draft of the manuscript.

## Data Sharing Statement

De-identified categorized data of the participants can be provided on request basis after approval of ethics committee of the institute/organisation to which the requesting person belongs, stating the potential need and usage and signing of an agreement with the corresponding authors on behalf of CSIR-IGIB. The request could be made to the corresponding authors after 90 days of publication.

## Acknowledgment

SSG would like to acknowledge CSIR grant MLP 2007 for this work. SN, AKB and RajatU would like to thank CSIR for their fellowships. SPrakash would like to acknowledge CSIR grant MLP-2002(CSIR-IGIB) for the fellowship. NB would like to address CSIR grant GAP-0192(CSIR-IGIB) for this work. We would also like to thank Directors of all the CSIR Institutes for facilitating the study. The authors also thank; Pushpesh Ranjan, Jitendra, Neeraj Kumar, Abhijeet and Rajkumar from AMPRI. V.Santosh Kumar from CCMB. Dr. Chandra Prakash Pandey from CDRI. Vipul Sharma, Akansh Agarwal, Hansraj Choudhary, Vijay Chatterjee, Narendra Meena, Ved Prakash, Alok Mishra, Navin Singhal, Ankit Shukla and Sudeep Rathore from CEERI. Avilash S Rani C and Naveen Shashidhar Kumbar from CFTRI. Swachchha Majumdar from CGCRI. Dr Dnyaneshwar Umrao Bawankule, Dr Debabrata Chanda, Pankaj Shukla, Sanjay Singh, Dr Dayanandan Mani, Ravi Kumar, Pankaj Yadav, Parmanand Kumar from CIMAP. D C Sharma, Dr Neelam J Gupta, A K Jain and Sudhansu Bhagat from CRRI. Pankaj Pandey, Rajesh, Dr. Mohammed Faruq and Ajay Pratap Singh from IGIB. Yogita Singh and Karvan Kaushal from CSIO. Jaykumar Patel, Shrikant Khandare and Dr Kannan Srinivasan from CSMCRI. Dr. Prakash L, Ganapathi G Bhatt, Shashikala U, Shashidhar K N from NAL. Dr. Vidyadhar Mudkavi, Ravichandran C and Sunil Babu M G from 4PI. Dr. Robin Singh, Mahesh S, Mohit Kumar Swarnakar, Dr. Pankaj Kulurkar from IHBT. Saikat Chakrabarti and Sandip Paul from IICB. Siva Ranjith and B Vijay Kumar from IICT. Sajad Ahmed from IIIM. Rene Christina, Neha Bansal and Ayan Banerjee from IIP. S.K. Goyal from NEERI-Delhi. Dr. P.R Meganathan and Dr. Shaik Basha from NEERI-Hyderabad.

Antara Sharma from NEIST. Dr. Shuchismita Benzwal and Chaitanya Dinesh from NGRI. Shana S Nair from NIIST. N. Anandavalli and P. Vasudevan from SERC. Vibha Malhotra Swaney from TKDL. Rashmi Arya and Prafulla Malwadkar from URDIP. Ajeet Singh and Dr. R.K.Sinha from HRDC.

## References

1. https://www.who.int/director-general/speeches/detail/who-director-general-s-opening-remarks-at-the-media-briefing-on-covid-1911-march-2020.

2. Nishiura H, Kobayashi T, Miyama T, et al. Estimation of the asymptomatic ratio of novel coronavirus infections (COVID-19). Int J Infect Dis 2020; 94: 154–5.

3. Bai Y, Yao L, Wei T, et al. Presumed Asymptomatic Carrier Transmission of COVID-19. JAMA 2020; 323(14): 1406–7.

4. Brown TS, Walensky RP. Serosurveillance and the COVID-19 Epidemic in the US: Undetected, Uncertain, and Out of Control. JAMA 2020; 324(8): 749–51.

5. Oran DP, Topol EJ. Prevalence of Asymptomatic SARS-CoV-2 Infection : A Narrative Review. Ann Intern Med 2020; 173(5): 362–7.

6. Andrews M, Areekal B, Rajesh K, et al. First confirmed case of COVID-19 infection in India: A case report. Indian Journal of Medical Research 2020; 151(5): 490–2.

7. Murhekar M, Bhatnagar T, Selvaraju S, et al. Prevalence of SARS-CoV-2 infection in India: Findings from the national serosurvey, May-June 2020. Indian Journal of Medical Research 2020; 152(1): 48–60.

8. Malani A, Shah D, Kang G, et al. Seroprevalence of SARS-CoV-2 in slums versus non-slums in Mumbai, India. The Lancet Global Health.

9. Ray A, Singh K, Chattopadhyay S, et al. Seroprevalence of anti-SARS-CoV-2 IgG antibodies in hospitalized patients at a tertiary referral center in North India. medRxiv 2020: 2020.08.22.20179937.

10. Satpati P, Sarangi SS, Gantait K, et al. Sero-surveillance (IgG) of SARS-CoV-2 among Asymptomatic General population of Paschim Medinipur District, West Bengal, India(Conducted during last week of July and 1st week of August 2020) - A Joint Venture of VRDL Lab (ICMR), Midnapore Medical College & Hospital & Department of Health and Family Welfare,Govt. of West Bengal, Paschim Medinipur. medRxiv 2020: 2020.09.12.20193219.

11. Murhekar M, Bhatnagar T, Selvaraju S, et al. SARS-CoV-2 Antibody Prevalence in India: Findings from the Second Nationwide Household Serosurvey, August - September 2020. SSRN; 2020.

12. Dhand NK, & Khatkar, M. S. Statulator: An online statistical calculator. Sample Size Calculator for Estimating a Single Proportion. 2014. http://statulator.com/SampleSize/ss1P.html.

13. Krüttgen A, Cornelissen CG, Dreher M, Hornef MW, Imöhl M, Kleines M. Determination of SARS-CoV-2 antibodies with assays from Diasorin, Roche and IDvet. Journal of virological methods 2021; 287: 113978.

14. Patidar GK, Dhiman Y. Distribution of ABO and Rh (D) Blood groups in India: A systematic review. ISBT Science Series; n/a(n/a).

15. Mohan P, Lando HA, Panneer S. Assessment of Tobacco Consumption and Control in India. Indian Journal of Clinical Medicine 2018; 9: 1179916118759289.

16. Koopmans M, Haagmans B. Assessing the extent of SARS-CoV-2 circulation through serological studies. Nature medicine 2020; 26(8): 1171–2.

17. Ortolan A, Lorenzin M, Felicetti M, Doria A, Ramonda R. Does gender influence clinical expression and disease outcomes in COVID-19? A systematic review and meta-analysis. International Journal of Infectious Diseases 2020; 99: 496–504.

18. Zhao J, Yang Y, Huang H, et al. Relationship between the ABO Blood Group and the COVID-19 Susceptibility. medRxiv 2020: 2020.03.11.20031096.

19. Wu Y, Feng Z, Li P, Yu Q. Relationship between ABO blood group distribution and clinical characteristics in patients with COVID-19. Clinica Chimica Acta 2020; 509: 220–3.

20. Göker H, Aladağ Karakulak E, Demiroğlu H, et al. The effects of blood group types on the risk of COVID-19 infection and its clinical outcome. Turk J Med Sci 2020; 50(4): 679–83.

21. Severe Covid GG, Ellinghaus D, Degenhardt F, et al. Genomewide Association Study of Severe Covid-19 with Respiratory Failure. N Engl J Med 2020; 383(16): 1522–34.

22. Golinelli D, Boetto E, Maietti E, Fantini MP. The association between ABO blood group and SARS-CoV-2 infection: A meta-analysis. PLOS ONE 2020; 15(9): e0239508.

23. Padhi S, Suvankar S, Dash D, et al. ABO blood group system is associated with COVID-19 mortality: An epidemiological investigation in the Indian population. Transfus Clin Biol 2020; 27(4): 253–8.

24. Conte L, Toraldo DM. Targeting the gut–lung microbiota axis by means of a high-fibre diet and probiotics may have anti-inflammatory effects in COVID-19 infection. Therapeutic Advances in Respiratory Disease 2020; 14: 1753466620937170.

25. Trompette A, Gollwitzer ES, Yadava K, et al. Gut microbiota metabolism of dietary fiber influences allergic airway disease and hematopoiesis. Nature medicine 2014; 20(2): 159–66.

26. Jayawardena R, Sooriyaarachchi P, Chourdakis M, Jeewandara C, Ranasinghe P. Enhancing immunity in viral infections, with special emphasis on COVID-19: A review. Diabetes & metabolic syndrome 2020; 14(4): 367–82.

27. Fontanet A, Tondeur L, Madec Y, et al. Cluster of COVID-19 in northern France: A retrospective closed cohort study. medRxiv 2020: 2020.04.18.20071134.

28. Makoto Miyara FT, Valérie Pourcher, Capucine Morelot-Panzini, et al. Low rate of daily active tobacco smoking in patients with symptomatic COVID-19. Qeios.; 2020.

29. Rossato M, Russo L, Mazzocut S, Di Vincenzo A, Fioretto P, Vettor R. Current smoking is not associated with COVID-19. European Respiratory Journal 2020; 55(6): 2001290.

30. Petrilli CM, Jones SA, Yang J, et al. Factors associated with hospital admission and critical illness among 5279 people with coronavirus disease 2019 in New York City: prospective cohort study. BMJ 2020; 369: m1966–m.

31. Farsalinos K, Barbouni A, Niaura R. Systematic review of the prevalence of current smoking among hospitalized COVID-19 patients in China: could nicotine be a therapeutic option? Intern Emerg Med 2020; 15(5): 845–52.

32. Simons D, Shahab L, Brown J, Perski O. The association of smoking status with SARS-CoV-2 infection, hospitalization and mortality from COVID-19: a living rapid evidence review with Bayesian meta-analyses (version 7). Addiction; n/a(n/a).

33. Polverino F. Cigarette Smoking and COVID-19: A Complex Interaction. Am J Respir Crit Care Med 2020; 202(3): 471–2.

34. Cai G, Bossé Y, Xiao F, Kheradmand F, Amos CI. Tobacco Smoking Increases the Lung Gene Expression of ACE2, the Receptor of SARS-CoV-2. Am J Respir Crit Care Med 2020; 201(12): 1557–9.

35. Oakes JM, Fuchs RM, Gardner JD, Lazartigues E, Yue X. Nicotine and the renin-angiotensin system. Am J Physiol Regul Integr Comp Physiol 2018; 315(5): R895–R906.

36. Figueiredo-Campos P, Blankenhaus B, Mota C, et al. Seroprevalence of anti-SARS-CoV-2 antibodies in COVID-19 patients and healthy volunteers up to 6 months post disease onset. European Journal of Immunology; n/a(n/a).

37. Seow J, Graham C, Merrick B, et al. Longitudinal observation and decline of neutralizing antibody responses in the three months following SARS-CoV-2 infection in humans. Nature Microbiology 2020; 5(12): 1598–607.

38. Deshpande G, Sapkal G, Tilekar B, et al. Neutralizing antibody responses to SARS-CoV-2 in COVID-19 patients. Indian Journal of Medical Research 2020; 152(1): 82–7.

